# COVID-19 Progression Timeline and Effectiveness of Response-to-Spread Interventions across the United States

**DOI:** 10.1101/2020.03.17.20037770

**Authors:** Pai Liu, Payton Beeler, Rajan K. Chakrabarty

## Abstract

Motivated by the rapid upsurge of COVID-19 cases in the United States beginning March 2020, we forecast the disease spread and assess the effectiveness of containment strategies by using an estalished network-driven epidemic dynamic model. Our model is initialized using the daily counts of active and confirmed COVID-19 cases across the US. Based on our model predictions for the March 14-16 timeframe, the national epidemic peak could be expected to arrive by early June, corresponding to a daily active count of ≈ 7% of the US population, if no containment plans are implemented. Epidemic peaks are expected to arrive in the states of Washington and New York by May 21 and 25, respectively. With a modest 25% reduction in COVID-19 transmissibility via community-level interventions, the epidemic progression could be delayed by up to 34 days. Wholesale interstate traffic restriction is ineffective in delaying the epidemic outbreak, but it does desynchronize the arrival of state-wise epidemic peaks, which could potentially alleviate the burden on limited available medical resources. In addition to forecasting the arrival timeline of the state-wise epidemic peaks, we attempt at informing the optimal timing necessary to enforce community-level interventions. Our findings underscore the pressing need for preparedness and timely interventions in states with a large fraction of the vulnerable uninsured and liquid-asset-poverty populations.

**Forecast website:** https://sites.google.com/view/covid19forecast

## Main Text

In December 2019, a novel coronavirus named SARS-CoV-2 began infecting residents of Wuhan, China (*1-3*). SARS-CoV-2 causes moderate to severe respiratory symptoms that can progress to severe pneumonia (coronavirus disease 2019, COVID-19) with an overall case-fatality rate of 2.3%, with 49.0% of cases becoming critical (*4*). Despite the quick responses and extreme disease containment measures taken in China (*5*), COVID-19 has spread rapidly to numerous countries and evolved into a global pandemic (*1, 3*). On January 30, 2020, the World Health Organization declared a “public health emergency of international concern” (*6*), and on the following day the United States Department of Health and Human Services declared a public health state of emergency (*7*).

During the week of February 23, 2020 the US Centers for Disease Control (CDC) reported new confirmed cases of COVID-19 in California, Oregon, and Washington, indicating the onset of “community spread” across the US (*7*). Until March 2, the total number of confirmed active COVID-19 cases in the US were 33, with new cases emerging in states of Texas, Arizona, Wisconsin, Illinois, Florida, New York, Rhode Island, and Massachuseets (*8*). In the following two weeks, this number has rapidly increased to 527 confirmed cases on March 9, and then to 4,216 cases on March 16; Altogether 49 US states, along with Washington DC and Puerto Rico, have already been seeded with COVID-19 patients (*8*). State of California and New York have respectively declared state emergency on March 4 and 7 (*9, 10*). The White House declared national emergency on March 13 (*11*). Thus, major outbreak of COVID-19 epidemic across the US is inevitable.

Here, we forecast the spread of COVID-19 in the US beginning March 2020 using a network-driven epidemic dynamics model that accounts for domestic interstate mobility across the country (Figure 1). The effectiveness of disease transmissibility reduction and interstate traffic restrictions are individually investigated. Risks posed by COVID-19 are analyzed state-wise with respect to the parameters: urgency, local uninsured populations, liquid assets poverty rate, and age weighted mortality rate.

**Figure 1.**
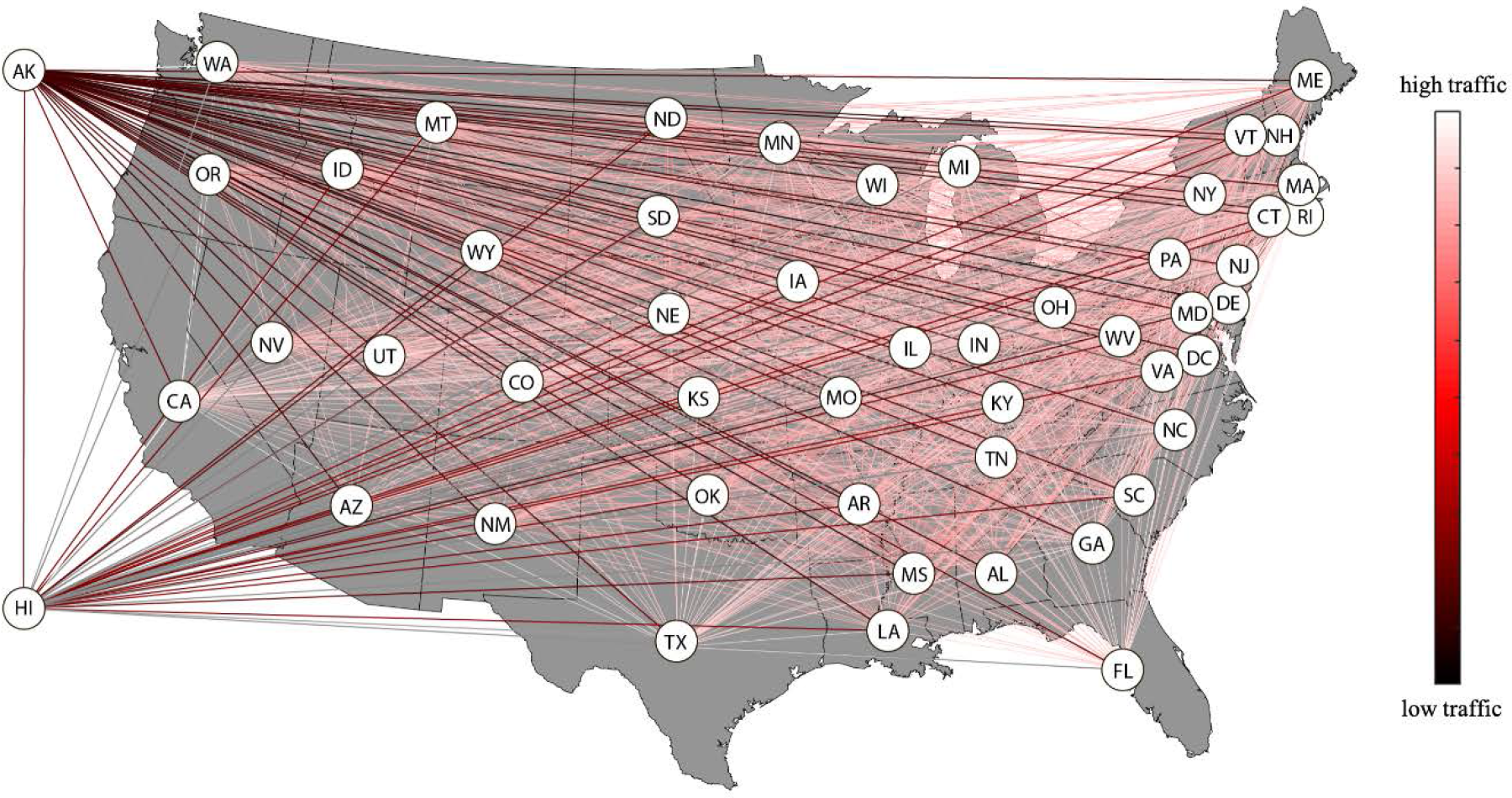
US domestic air traffic pattern generated with the 2018-2019 aviation statistics and state-wise population. Segiments are colored according to the air traffic volume (Detailed in Supplementary Materials (*19*)) between the corresponding states.

### Modelling the Network-Driven Epidemic Dyamics of COVID-19

We simulated the COVID-19 epidemic spread in the United States using a Susceptible-Exposed-Infected-Recovered (SEIR) model (*1, 2, 12, 13*) coupled with network-driven dynamics (*14-16*) accounting for the domestic air traffic taking place amongst the 50 US states, Washington DC, and Puerto Rico (hereafter generically denoted as states). In the SEIR model, fractions of susceptible (*s*_*n*_), exposed (*e*_*n*_), infected (*j*_*n*_), and recovered (*r*_*n*_) individuals are tracked within a state *nn* per the kinetics of mass action; The interstate exchange of passengers is captured with a matrix of *P*_*mn*_ quantifying the probability that an individual leaving state *nn* ends up in *m* (*14*). The governing equations can be expressed as a set of first-order differential equations with respect to time (*t*):

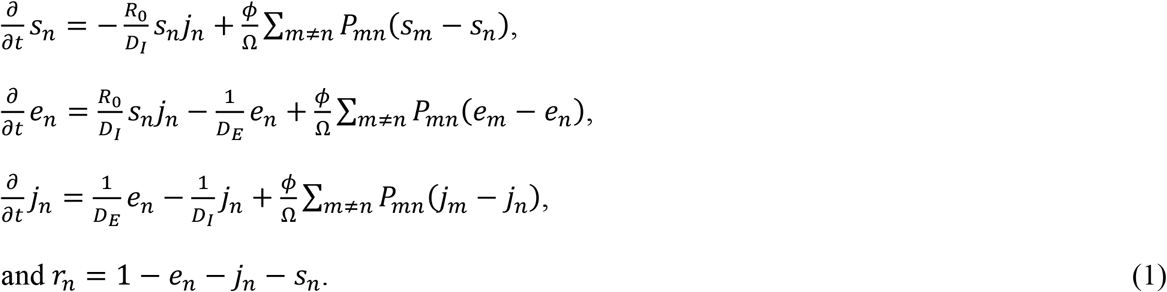

Here, *R*_0_, *D*_*E*_, and *D*_*I*_ represent the basic reproduction ratio, mean incubation period, and mean infectious period, respectively, of COVID-19 (*1, 2, 17*); *ϕ* is the daily passenger flux of the entire air traffic network, and Ω represents the total US population (*14, 18*). The ratio *ϕ*/Ω can be regarded as an interstate mobility parameter. Equation (1) was integrated numerically with a discrete *t* that increment in units of one day. Our integration involved dynamically updating *ϕ* and *P*_*mn*_, which are monthly resolved dataset, per changing *t*. The resulting *j*_*n*_(*t*) was next used to calculate the fraction of infected population on a national level, *j*_*natl*_(*t*):

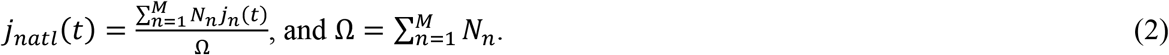

Here, *N*_*n*_ is the state-wise population data, and *M*=52 is the total number of locale (50 states, Washington DC, and Puerto Rico). It was assumed in our model that the interstate exchange of passengers is predominately via air traffic since volume of ground-based exchange is negligible (see Supplementary Materials (*19*)). We further assumed that the long term international importation of individual infected with COVID-19 is minimal, under the travel restrictions enforced on international passengers that arrive from countries and regions where COVID-19 is widespread (*20,21, 22*). The control of COVID-19 transmissibility and interstate mobility were realized by adjusting the values of *R*_0_ and *ϕ*, respectively (*2*). For example, a 25% reduction in transmissibility of COVID-19 can be built in the model by substituting *R*_0_ in Eq. (1) with *R*_0_^’^ = 0.75*R*_0_. Similar treatment was done on *ϕ* when the influence of reduced interstate mobility on epidemic dynamics was investigated.

Forecasts have been performed in the timeframe between March 2 and 16. For each forecast, the SEIR model was initialized with the daily confirmed active cases of COVID-19 in the US (Refer to Table S2 in Supplementary materials (*19*)), which were acquired from a web-based dashboard for real-time epidemic tracking published by Johns Hopkins University (*8*). The matrices of *P*_*mn*_(hereafter **P**) and *ϕ* were calculated using the latest monthly aviation data (between September 2018 and August 2019) released by the United States Bureau of Transportation Statistics (*23*) (The calculation of **P** and *ϕ* is detailed in Supplementary Materials (*19*)). The state-wise population *N*_*nn*_ was acquired from database of United States Census Bureau (*24*). The *R*_0_, *D*_*E*_, and *D*_*I*_ of COVID-19 were assumed to be 2.68, 6 days, and 2.4 days, respectively, per the values recommended in Ref. (*2*).

### The COVID-19 Epidemic Dynamics Without Intervention

Figure 2 shows forecast of SEIR model initialized on March 16 under a status quo condition. Panel (a) shows the dynamics of COVID-19 spread in the US, highlighting the temporal evolution of infected population *j*_*n*_(*t*) state-by-state sorted by the earliness of the arrival of local epidemic peak. Panel (b) maps the spread pattern of COVID-19 by showing a time series of continental US map wherein each state is colored according to the local infected fraction. Among the earliest outbreaks along the pacific west region, epidemics in Washington will reach a local peak by May 21. Immediately followed is the state of New York, wherein a local epidemic peak is expected to arrive by May 25. Alaska will experience a rapid increase in local infected fraction, which will peak around May 29, because of its frequent air traffic commuting from and to Washington. Similar trend is observed for Hawaii, where the local epidemic is estimated to peak on June 3, under the influence of high passenger flux emanating from California whose local epidemic peak will arrive around June 1. The local epidemics in Massachusetts, Washington DC, Colorado, and New Jersey will peak by the end of May, which is followed by Illinois, Georgia, and Florida, where the local epidemic peaks are expected to arrive by early June. The majority of states will experience epidemic peaks during the first week of June. The daily active COVID-19 cases at the epidemic peak (hereafter, epidemic peak magnitude), is uniform across each state with a median value ≈ 7.59% of the state-wise population. The national epidemic peak is estimated to arrive on June 3, with an daily active cases ≈ 7.16% of total US population.

**Figure 2.**
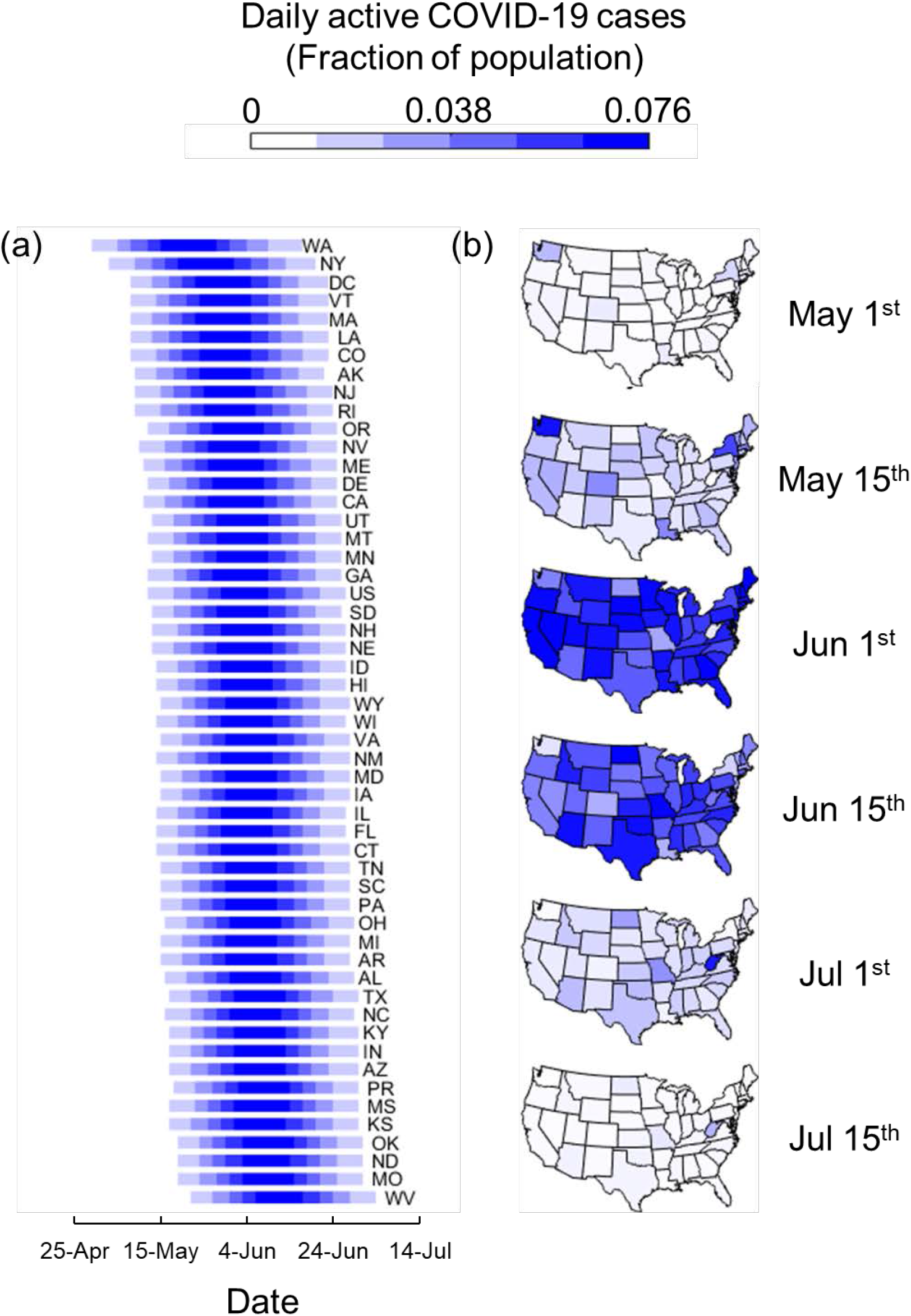
Forecasting the spread dynamics of COVID-19 epidemics in the US. **(a)** Epidemic forecast for 50 states in the US, along with Washington DC and Puerto Rico. Forecast is made under baseline condition that no control on transmissibility and interstate traffic is implemented. **(b)** Map of the continental US colored according to the state-wise epidemic magnitude over the period between May and August 2020.

### Intervention Effectiveness

Figure 3 shows the influences of interstate mobility reduction and disease transmissibility reduction on epidemic dynamics. These influences are quantitated with respect to two factors – the delay of epidemic peak (panel (a) and (c)) and the reduction in peak magnitude (panel (b) and (d)). In these series of plots, the forecasts of SEIR model initialized on March 2 (symbols in red) and March 16 (Symbols in black) are overlaid, with box-and-whiskers and circles representing state-wise and national statistics, respectively.

**Figure 3.**
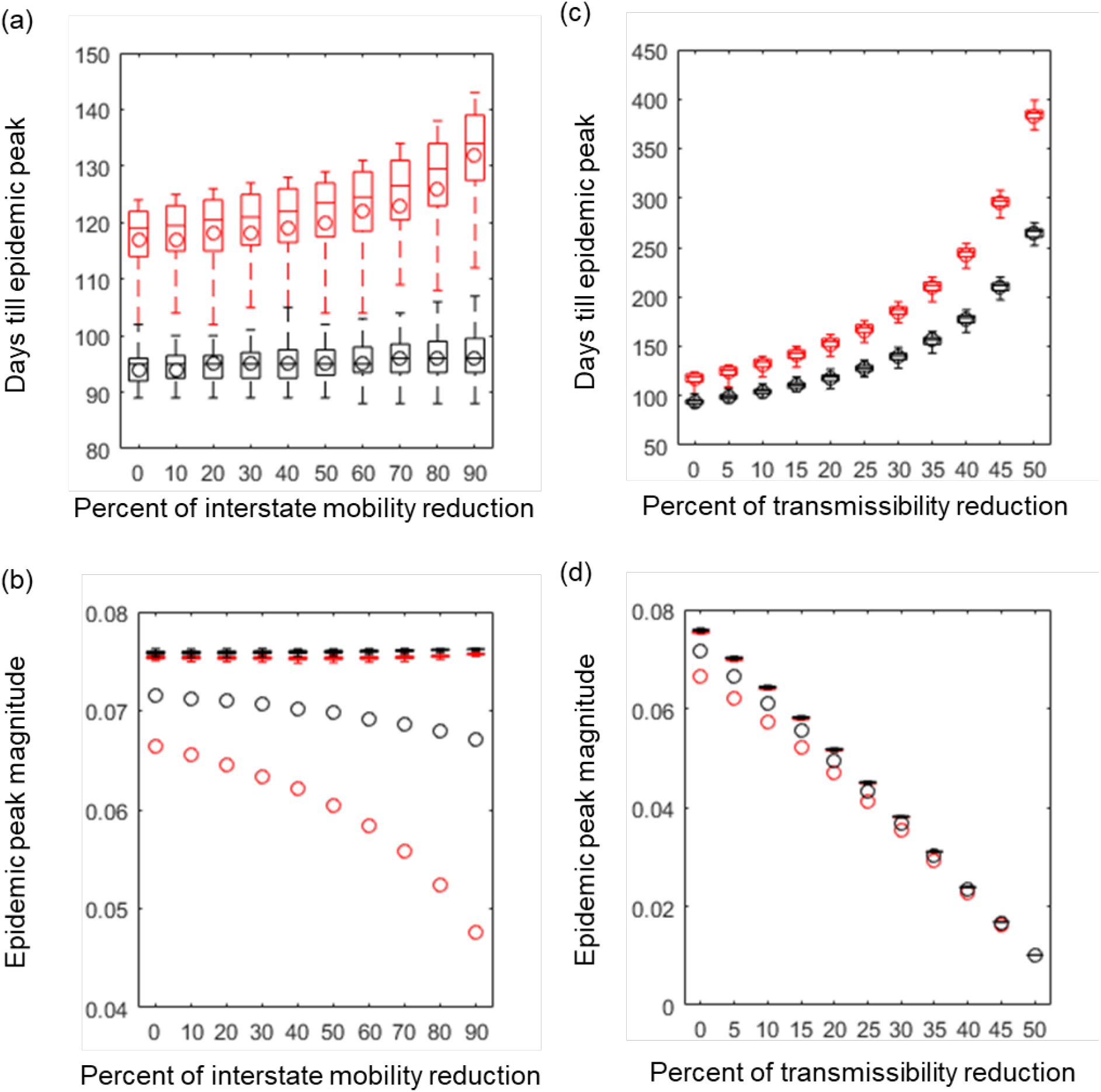
Effectiveness of Response-to-Spread Interventions. Circles represent national statistics; Black color represents the forecast of SEIR model initialized on March 16; Red color represents that initialized on March 2) **(a)** Epidemic peak arrival time (measured in days starting March 1) as a function of percent of interstate mobility reduction **(b)** Epidemic peak magnitude as a function of interstate mobility reduction. **(c)** Epidemic peak arrival time as a function of percent of transmissibility reduction. **(d)** Epidemic peak magnitude as a function of transmissibility reduction. (Boxes and whiskers represent state-wise statistics; Circles represent national statistics; Black color represents the forecast of SEIR model initialized on March 16; Red color represents that initialized on March 2)

Figure 3 (a) indicates that it is already too late (up to now, March 16, black symbols) for a wholesale traffic restriction to be effective in containing the epidemic spread, since virtually all US states have been seeded with COVID-19 patients. The previous modelling study on the COVID-19 spread in mainland China (*2*) has also arrived at a similar conclusion regarding the futility of mobility reduction in delaying epidemic spread. Nonetheless, it is worth noting that the reduction in interstate mobility in the US could slightly desynchronize the state-wise epidemic dynamics (i.e. the moderate broadening of the time window for the local epidemic peaks shown in panel (a)). This desynchronization could potentially alleviate the burnden on the limited available medical resources on a national level. The side-by-side comparison with the forecast made on March 2 (red symbols) indicates that should an aggressive (c.a. 90% mobility reduction) traffic restriction have been emplemented two weeks before, the national epidemic peak could have been delayed substaintially. Panel (b) shows that 90% reduction of interstate mobility, if implemented on March 2, could have reduced the epidemic peak magnitude by up to 30%. However, up to March 16, the influence of mobility reduction on epidemic peak magnitude has dwindled to a negligible level.

Figure 3 (c) and (d) show that till date reducing COVID-19 transmissibility remains to be an effective intervention approach. A 25% reduction in transmissibility (*R*_0_ reduced to 2.01) across all states could delay the national epidemic peak by about 35 days and reduce its magnitude by 39%. A 50% reduction in transmissibility (*R*_0_ reduced to 1.34) will contain the spread of COVID-19 (with the national epidemic peak postponed to winter 2020 and the peak magnitude reduced to 1%). These observations agree with Ref. (*2*) wherein a forecast of epidemic dynamics in China was made. The influence of transmissibility reduction on state-wise epidemic dynamics is uniform. Reduction in disease transmissibility can be achieved via a variety of control measures. For example, individual level measures include practices such as improving personal hygiene and maintaining social distance. Studies of past epidemics have shown that individual-scale control measures as simple as hand washing can reduce the risk of non-specific respiratory infection by 6%-44%, and was an effective method to control the transmissibility of SARS (*25, 26*). Another frequently adopted individual level practice is the use of masks, however, till date no consensus has been reached on its effectiveness (*27*-*29*). Community level interventions include strategies such as contact tracing and quarantine, business and school closure, and restricting mass public gatherings.

Figure 4 retrospectively shows the decay of intervention effectiveness over time. Panel (a) shows that the amount of time we can buy (with interstate mobility reduction and transmissibility reduction) decreases gradually and steadily during the first two weeks of March. Panel (b) benchmarks the interventions with an effectiveness factor – defined as the normalized percent of reduction in epidemic peak magnitude. The effectiveness of 90% interstate mobility reduction rapidly decreases to a negligible level; Whereas the effectiveness of 25% transmissibility remains constant. Again, we emphasize here that the opportunity window of containing COVID-19 via aggressive traffic restriction has already been closed, and therefore the limited resources should be directed towards reducing COVID-19 transmissibility on individual and community level.

**Figure 4.**
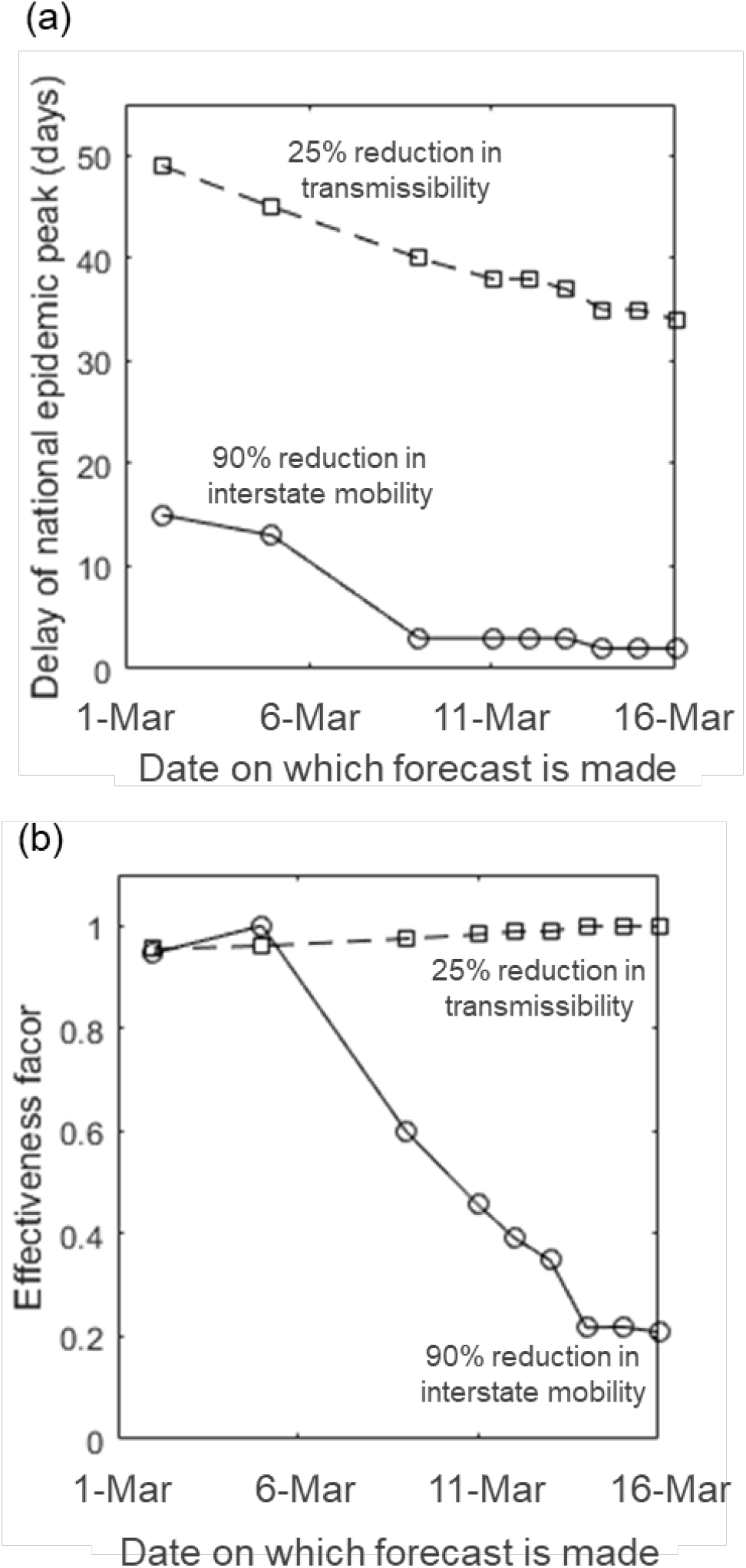
Intervention effectiveness decays over time. **(a)** Delay of national epidemical peak with 25% transmissibility reduction or 90% interstate mobility reduction, as a function of the date on which forecasts are made. **(b)** Effectiveness of interventions (quantified by normalized percent of reduction in epidemic peak magnitude) as a function of forecast date.

### Timing to Implement Finite Time Intervention

Previous modeling studies have shown that community level control measures are effective in reducing disease transmissibility, however, prolonged closures of schools and businesses, as well as limited public gatherings have negative socio-economic impacts that must be considered (*30, 31*). The negative socio-economic impacts dictate that the community-level interventions can only last a finite amount of time. Therefore, it is of utmost importance to understand *when* and for *how long* those interventions should be put into effect, so as to maximize the net benefit. Figure 5(a) shows the output of the SEIR model under a conditional 25% reduction in transmissibility within a time window defined with an intervention start time, *t*_*i*_, and an end time, *t*_*i*_ *+ τ* (where *ττ* represents intervention duration). Effectiveness of interventions (lasting for a variable *ττ*), which is quantified with the normalized reduction of infected population at the national epidemic peak, is plotted in Fig. 5 (a) as a function of implementation timing, which is quantified with number of weeks between *t*_*i*_ and the national epidemic peak. The trends in Fig. 5 (a) show that unless the intervention could last indefinitely, a premature implementation of a finite-time intervention plan could be counterproductive. Figure 5 (b) plots the best implementation timing, *t*^*∗*^, as a function of *τ*, and a power-law relationship *t*^*∗*^ ≈ *τ*^0.85^ is conceived. This empirical relationship informs the optimal timing to enforce community-level interventions given a practically affordable intervention duration. The earliness of the state-wise epidemic peaks are presented in Fig. 5 (c).

**Figure 5.**
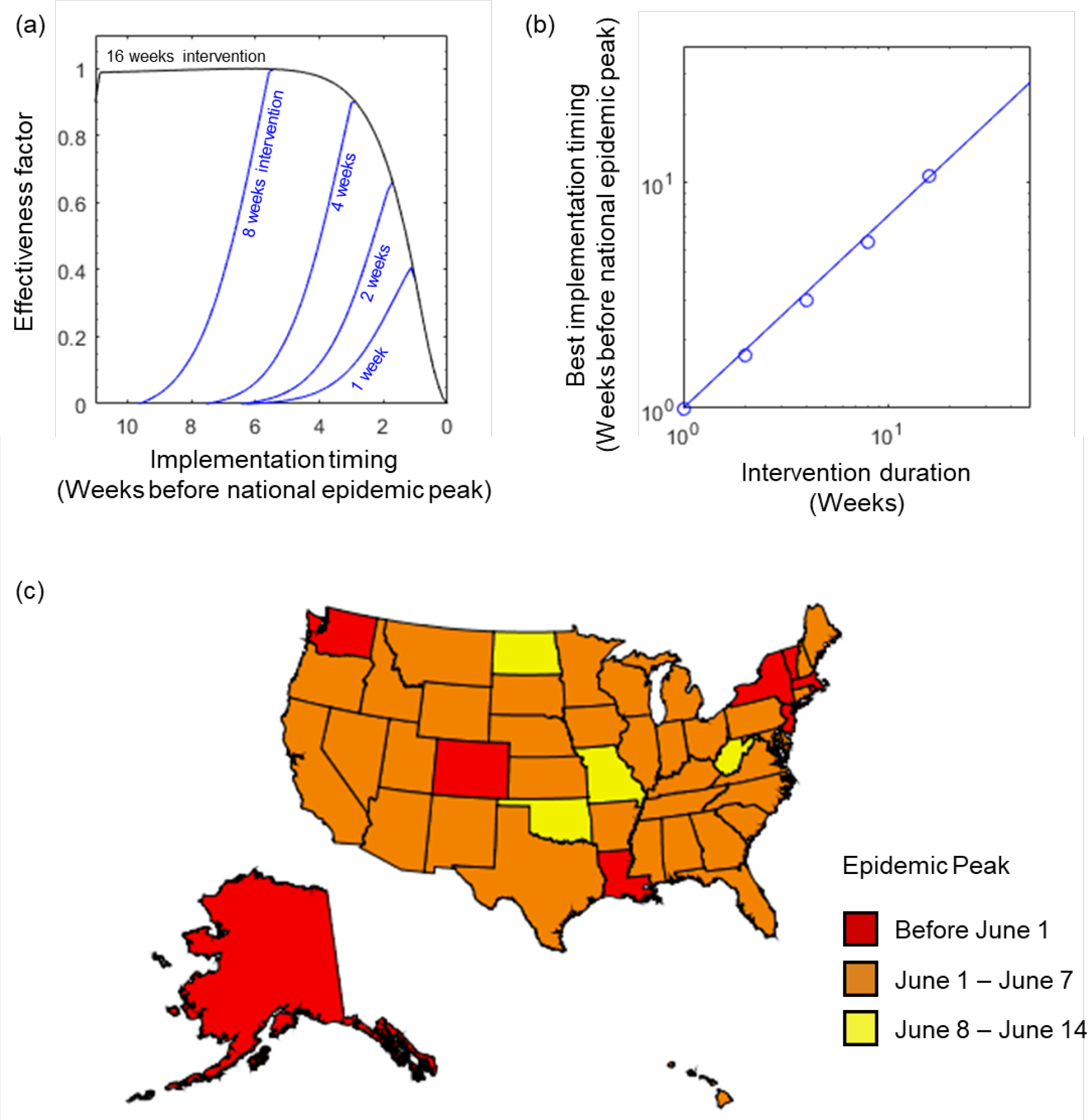
Timing and effectiveness of the intervention targeted on reducing virus transmissibility. **(a)** Effectiveness (quantified by normalized percent of reduction in epidemic peak magnitude) as a function of the implementation timing (quantified with number of weeks ahead of national epidemic peak). **(b)** Empirical relationship between the best implementation timing and intervention duration. Solid line represents a power-law fit with an exponent of 0.85. **(c)** Map of the US with each state clustered and colored per the earliness of local epidemic peak.

### State-wise Risk Assessment

Figure 6 shows an assessment of the vulnerability of each state against COVID-19 epidemics. Our assessment was done on four core factors, (a) urgency, (b) percent of uninsured local population, (c) local liquid-asset-poverty rate, and (d) age-weighted mortality rate of the infected patients. Urgency is quantitated as the date of a local epidemic peak per the SEIR model; it identifies how much time is left for the local policy makers and disease control professionals to design and implement effective interventions. The percent of uninsured population is negatively related to the overall willingness of the local resident to seek out diagnosis and quarantine as and when the early symptoms of COVID-19 arise (*32*). The percent of liquid-asset-poverty is the fraction of local households that lack savings to sustain above poverty level for three-months (*33*). The implication of lengthy societal shutdown is catastrophical to these households or individuals because of the potential disruption in their income. The age-weighted mortality rate quantifies the magnitude of consequence should every intervention on disease containment fail and the situation evolve to disrest. A summary report (*4*) of the COVID-19 cases by Chinese Center for Disease Control and Prevention has identified that the case-fatality rate strongly correlates with patients’ age. The overall fatality rate for all confirmed cases in China is 2.3% (1023 of 44672 cases) (*4*); however, patients aged ≥ 80 years and 70-79 years have case fatality rate of 14.8% (208 of 1408 cases) and 8.0% (312 of 3918 cases), respectively. In our analysis, the age-weighted mortality rate for a state was calculated using age-specific fatality rate reported in Ref. (*4*) in conjunction with state-wise demographic composition (*19*).

**Figure 6.**
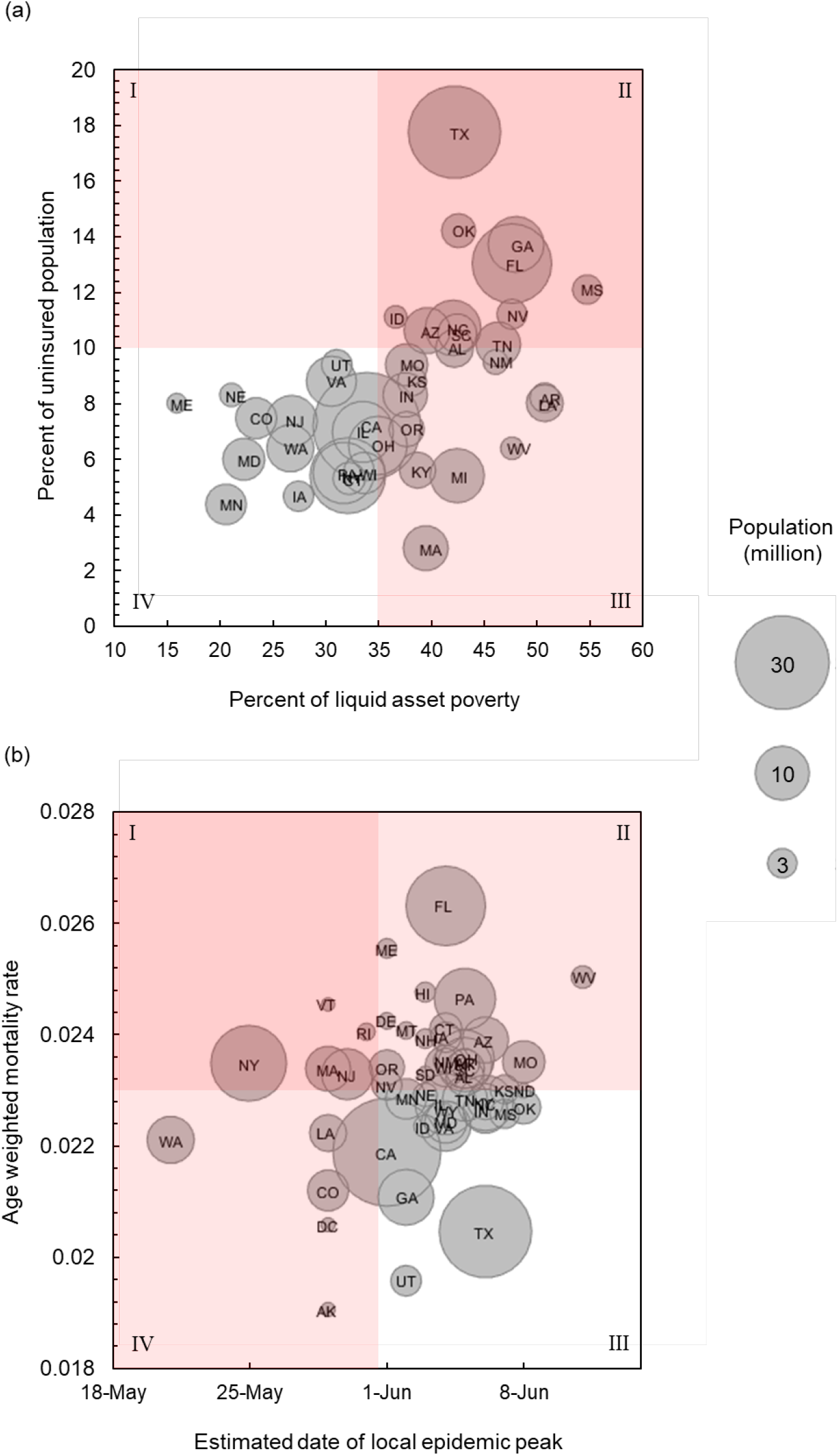
State-wise risk assessment. **(a)** States are clustered according to the state-wise liquid-asset-poverty rate and state-wise uninsured population. **(b)** States are clustered according to the earliness local epidemic peak and age-averaged mortality rate. The size of the circle symbol is proportional to the state-wise population. Red color shading indicates severity.

Figure 6 (a) shows how each state is expected to be affected with regard to the percent of uninsured population and the liquid-asset-poverty rate. A positive correlation can be observed in the dataset, since the individuals who live on paycheck-by-paycheck are less likely to be enrolled in medical insurance. The states scattered in qudrant II, such as Texas, Georgia, Oklahoma, Florida, and Mississippi, are vulnerable to the impact of COVID-19 due to their high liquid asset poverty rate. Among these states, Texas is especially notable because of the sheer amount of uninsured population. Figure 6 (b) plots each state according to date of local epidemic peak and the age-weighted mortality rate. The situation in Washington is considerably urgent, followed by New York, Washington DC, Massachusetts, Alaska, Louisiana, Colorado, and Vermont wherein the local epidemic peaks are expected to strike much earlier compared to the rest of the country. States with large populations, such as Florida, Pennsylvania, Arizona, and New York, would face an above-average mortality rates because of their higher degree of local population aging.

### Conclusion

In this work, we performed a modelling study on the COVID-19 epidemic spread across the US using the epidemiological parameters observed from China. Our prognosis suggests that in the absence of disease control interventions and traffic restrictions, the nation-wide infected fraction could reach ∼7% when the epidemic hits its peak by early June. If the transmissibility of COVID is reduced by 25% with respect to its baseline, the national epidemic peak could be delayed by about 34 days, along with a 39% reduction in peak magnitude. The COVID-19 epidemic in the US would fade out if a 50% reduction in disease transmissibility could be achieved across all states. Interstate mobility reduction is shown to be ineffective on delaying epidemic outbreak, but a wholesale interstate traffic restrictions slightly desynchronizes the arrival of state-wise local epidemic peaks, which could potentially alleviate burdens on medical resources. The timing of implementation of large scale disease control intervention is crucial, especially when the local population cannot withstand a lengthy society shutdown. State-by-state analysis shows that the state of Washington, New York, Washington DC, Massachusetts, Alaska, Louisiana, Colorado, and Vermont could be impacted by an earlier arrival of local epidemic peaks than the rest of the country. Texas, Georgia, Oklahoma, and Florida face the risks posed by their large uninsured population and high liquid asset poverty rate. Florida, Pennsylvania, Arizona, and Florida share a higher overall COVID-19 mortality rate due to their aged demographic composition.

## Data Availability

All Processed data are made available in the Supplementary Materials of the manuscript and online via the website: https://sites.google.com/view/covid19forecast

